# Coronavirus infection in neonates: Neurodevelopmental outcomes at 18 months of age

**DOI:** 10.1101/2022.04.15.22273460

**Authors:** Mariam Ayed, Zainab Alsaffar, Zainab Bahzad, Yasmeen Buhamad, Ali Abdulkareem, Alaa AlQattan, Alia Embaireeg, Mais Kartam, Hessa Alkandari

## Abstract

**Background:** Although most neonates with severe acute respiratory syndrome coronavirus-2 (SARS-CoV-2) infections have mild disease, the impact on neurodevelopmental outcomes is unknown. This study aimed to assess 18-month neurodevelopmental outcomes of neonates infected with SARS-CoV-2 infection.

**Methods:** We conducted a prospective cohort study of neonates diagnosed with SARS-CoV-2 infection between June 2020–August 2020 through nasopharyngeal coronavirus disease 2019 (COVID-19) PCR testing. A total of 58 neonates were identified from the Kuwait national COVID-19 registry and were enrolled. Historical controls were selected from the neonatal follow-up registry and matched 2:1 based on sex and gestational age. At 18 months of age, neurodevelopmental outcomes were assessed using Bayley Scales of Infant and Toddler Development-3rd Edition (BSID-III) by two trained assessors.

**Results:** A total of 40 children were diagnosed with SARS-CoV-2 infection and included in the final analysis. The median age at infection was 18 days (range: 10-26 days). Eighteen (45%) were asymptomatic, 15 (37.5%) had a sepsis-like presentation, 5 (12.5%) had respiratory distress and 2 (5%) had a multisystem inflammatory syndrome in children (MIS-C)-like presentation. At 18 months follow up, only one child had severe developmental delay, and one child had a language delay. BSID-III outcomes did not significantly differ between the SARS-CoV-2 infected group and the control group.

**Conclusions:** There is no difference in neurodevelopmental outcomes in children infected with SARS-CoV-2 infection at 18 months compared to controls, although longer neurodevelopmental follow-up studies are required.

## 1. INTRODUCTION

Severe acute respiratory syndrome coronavirus-2 (SARS-CoV-2) infection in children has been associated with a multisystem inflammatory syndrome and coronavirus disease 2019 (COVID-19), and these are recognized as two different clinical entities [1-3]. It is widely known that, overall, a smaller proportion of children become ill when infected with SARS-CoV-2 compared to adults [4].

Neurological symptoms have been identified in paediatric cases of COVID-19 [5], with approximately 22% of hospitalized children being affected by neurological manifestations such as smell/taste disorders, headache, and stroke [6]. Intracranial hypertension [7] and acute encephalitis [8] have also been reported in children with multisystem inflammatory syndrome.

There are a range of potential mechanisms through which CNS involvement may occur in association with COVID-19. Animal studies of SARS-CoV have suggested that the angiotensin-converting-enzyme-2 receptor may mediate coronavirus-related neuronal damage, and there is also evidence that the virus can infect the cerebrovascular endothelium and brain parenchyma, particularly in the medial temporal lobe, with resulting apoptosis and necrosis [9]. In addition, there is also evidence from human postmortem brain studies that human coronavirus variants and SARS-CoV can infect both neurons and glia, and increased cytokine serum levels associated with SARS-CoV infection may indicate increased cytokine production [9]. The presence of autoantibodies in the cerebrospinal fluid of patients with COVID-19 and neurological symptoms also raises the possibility of autoimmune mechanisms being involved in the pathogenesis of COVID-19 with neurological involvement [10].

Although most neonates with SARS-CoV-2 infections have mild disease, the impact on long-term neurodevelopmental outcomes is currently unknown. It has been shown that birth during the COVID-19 pandemic, but not in utero exposure to maternal SARS-CoV-2 infection, is associated with neurodevelopmental outcome differences at age 6 months [11]. Another study has shown that newborns exposed in utero to SARS-CoV-2 have largely normal neurological development in the first few months of life [12]. However, these studies provide only a short-term perspective on neurodevelopmental outcome.

The objective of this study was to assess the neurodevelopment outcomes at 18 months in neonates following confirmed SARS-CoV-2 infection. It was hypothesized that infection with SARS-CoV-2 virus may lead to adverse neurodevelopmental outcomes in neonates.

## 2. MATERIALS AND METHODS

We did a prospective cohort study of neonates diagnosed with SARS-CoV-2 infection between June 2020 and August 2020. Neonates undergo nasopharyngeal COVID-19 PCR according to our national COVID-19 protocol: 1) who had contact with a known COVID-19 patient 2) having symptoms of fever, cough or respiratory distress 3) screening test prior to hospital admission. Verbal informed consent obtained from the child’s parents prior to conducting the study. Ethics approval was granted by the Ministry of Health of Kuwait (approval number 2021-1638).

A total of 58 neonates were identified from the Kuwait national COVID-19 registry and were enrolled into the prospective SARS-CoV-2 infection outcome cohort. Of these, 6 children were lost to follow up, 2 refused to participate in the neurodevelopmental outcome study, 6 lift the country, one infant died from COVID-19 complication. Four neonates were unable to determine COVID-19 status due to equivocal result. Historical controls were selected from the neonatal follow up registry and matched 2:1 based on sex and gestational age.

Data were retrieved from hospital records and included information on demographic details, parental education, maternal age at the birth of the child, clinical features, hospital course and morbidity assessment including any seizure episode or rehospitalization.

We defined respiratory distress as the presence of one of the following: increased work of breathing or oximetry <93%. We used the multisystem inflammatory syndrome (MIS-C) definition of the CDC and AAP [13,14]. MIS-C often occurs in age <21 years with clinical criteria: A minimum 24-hour history of subjective or objective fever ≥38.0°C and severe illness necessitating hospitalization and 2 or more organ systems affected (i.e., cardiac, renal, respiratory, hematologic, gastrointestinal, dermatologic, neurological). Laboratory evidence of inflammation include 1 or more of the following: an elevated C-reactive protein (CRP), erythrocyte sedimentation rate, fibrinogen, procalcitonin, D-dimer, ferritin, lactate dehydrogenase, or interleukin-6; elevated neutrophils or reduced lymphocytes and low albumin. A diagnosis of central nervous system co-morbidities and chromosomal abnormalities were ruled out in the enrolled neonate.

At 18 months of age, neurodevelopmental outcomes using Bayley Scales of Infant and Toddler Development-3rd Edition (BSID-III) [15] were assessed by 2 trained assessors. The scales are age-standardized with the composite cognitive, motor and language scales having a normative mean of 100 and standard deviation (SD) 15. A moderate developmental delay was given for worst composite score of 70–84 in one or more of the 3 domains. Whereas, the severe developmental delay was used for a score was < 70 for any of the 3 domains, or when unable to assign a score owing to severe mental deficiency or cerebral palsy as appraised using the Gross Motor Function Classification System (GMFCS). Blindness and deafness were also assessed by a pediatrician. *Blindness* was defined as having visual acuity of less than 6/60 in the better eye, and *deafness* was defined as a hearing impairment requiring amplification or a cochlear implant or worse. We also obtained anthropometric parameters: head circumference, length, and weight measured utilizing standard techniques using growth standards by World Health Organization.

### 2.1 Statistical analysis

Categorical data were presented as proportions and continuous data were presented as median and interquartile range (IQR). Differences in clinical data between infants were compared with Pearson’s Chi square test for categorical variables (Fisher exact test was used when frequency was less than 5) and with Wilcoxon rank-sum test for continuous variables. P value <0.05 was considered statistically significant. Statistical analysis was performed with Stata/IC 14.2 (StataCorp, College station, Texas, 2015).

## 3. RESULTS

A total of 40 children were diagnosed with SARS-CoV-2 infection and included in the final analysis. The demographic details of the study cohort are shown in Table 1. The median age at infection was 18 days (IQR: 10–26 days). Two of the infected neonates were born to SARS-CoV-2 infected mothers at time of birth. Eighteen (45%) were asymptomatic, 15 (37.5%) had sepsis like presentation, 5 (12.5%) had respiratory distress, and 2 (5%) had MIS-C like presentation. Median duration of hospital stay was 4 days (IQR: 2–14 days).

**Table 1.** Demographic characteristics

| Variable | SARS-CoV-2 exposed<br>N=40 | Historical control<br>N= 80 | P value |
| --- | --- | --- | --- |
| Gestational age, week | 38 (37-40) | 39.2 (38-40) | 0.716 |
| Gender |  |  |  |
| Male | 19 (47.5%) | 39 (48.8%) | 0.872 |
| Female | 21 (52.5%) | 41 (51.2%) |  |
| Birth weight, grams | 3000 (2500-3420) | 3457 (3147-3765) | 0.12 |
| Mode of delivery |  |  | 0.532 |
| Caesarean section | 13 (57.5%) | 42 (52.5%) |  |
| Vaginal birth | 17 (42.5%) | 38 (47.5%) |  |
| 5 minutes Apgar score | 8 (7-10) | 8 (7-9) | 0.128 |
| Maternal education |  |  |  |
| Educated | 1 (2.5%) | 5 (6.2%) | 0.104 |
| Elementary education | 2 (5%) | 2 (4.5%) |  |
| High school | 4 (10%) | 7 (68.7%) |  |
| Diploma | 13 (32.5%) | 29 (36%) |  |
| Bachelor/PhD | 20 (50%) | 37 (46.2%) |  |
| Maternal age, years | 27.3 (25-32) | 27.7 (26-31.7) | 0.296 |
| Paternal education |  |  | 0.109 |
| Educated | 1 (2.5%) | 0 |  |
| Elementary education | 5 (12.5%) | 10 (12.5%) |  |
| High school | 4 (10%) | 12 (15%) |  |
| Diploma | 12 (30%) | 28 (35%) |  |
| Bachelor/PhD | 18 (45%) | 30 (37.5%) |  |
| Age at follow up, months | 18.6 (18.2-19.5) | 18.7 (18.4-19.3) | 0.174 |
| Data are expressed as Number (%) and median (interquartile range) |  |  |  |

During the 18 months follow up period, two infants required rehospitalization, one with simple febrile seizure and second with acute bronchiolitis.

No measured maternal or infant characteristics differed substantially between the exposed and unexposed group (Table 2).

**Table 2.** Follow up outcomes at 18 months

| Parameters | SARS-CoV-2 infected group<br>N= 40 | Control group<br>N= 80 | P value |
| --- | --- | --- | --- |
| Bayley-III |  |  |  |
| Motor composite score | 100 (91-107) | 97 (88-107) | 0.101 |
| 71-85 | 0 | 2 (2.5%) | 0.786 |
| <70 | 1 (2.5%) | 1 (1.2%) |  |
| Fine motor | 11 (10-12) | 10 (9-12) | 0.21 |
| Gross motor | 9 (7-10) | 9 (6-10) | 0.231 |
| Cognitive composite score | 105 (100-115) | 110 (100-115) | 0.952 |
| 71-85 | 0 | 3 (3.7%) | 0.261 |
| <70 | 1 (2.5%) | 0 |  |
| Language composite score | 100 (89-109) | 100 (89-109) | 0.785 |
| Expressive | 9 (7-12) | 9 (8-11) | 0.971 |
| Receptive | 10 (8-12) | 10 (8-12) | 0.645 |
| 71-85 | 1 (2.5%) | 3 (3.7%) |  |
| <70 | 1 (2.5%) | 1 (1.2%) | 0.864 |
| Cerebral palsy | 1 (2.5%) | 2 (2.5%) | 0.131 |
| Seizure | 1 (2.5%) | 0 | 1 |
| Deafness | 0 | 1 (1.2%) | 0.293 |
| Blindness | 0 | 0 |  |
| Weight (kg) | 11.1 (10.2-12.7) | 11.7 (10.7-12.9) | 0.212 |
| Length (cm) | 82 (79-84) | 83 (78-86) | 0.304 |
| Head Circumference (cm) | 47.6 (46.9-48.8) | 48.5 (47.5-49) | 0.138 |
| Data are expressed as number (%) or median, interquartile range (IQR) |  |  |  |

At 18 months, there were no differences in the Bayley-III composite scores between the exposed infants and the control group. In the SARS-CoV-2 exposed group, one infant had a score less than 70 in motor, cognitive and language composite scores and one infant had a language composite score less than 85. The infant with low scores <70 in all domain had neonatal MIS-C like presentation. She presented with lethargy, fever, tachycardia and hypotension. She required invasive ventilation, inotropic support and intravenous immunoglobulin. She developed attacks of seizures and had markedly increase in C-reactive protein, myocardial enzymes and D-Dimer. Lumbar puncture was not performed due to instability. MRI brain showed right temporal and bilateral basal ganglia hemorrhagic foci with cortical and subcortical abnormal signal intensity on T2-weighted images in both fronto-parietal and posterior parietal regions. The second child who scored less than 85 in language composite score, he presented with fever and pneumonia confirmed by chest radiographic image. He received supportive treatment for 5 days. He had normal hearing assessment and normal neurological exam.

## 4. DISCUSSION

This prospective cohort study compared the neurodevelopmental outcomes at 18 months in 40 children diagnosed with SARS-CoV-2 infection compared to a group of 80 historical controls that were well matched in terms of maternal and infant characteristics at baseline. The study found no significant difference in the Bayley-III composite scores between the two groups, indicating similar neurodevelopmental outcomes.

There has been much interest in the manifestations of SARS-CoV-2 infection in the pediatric population, including the neurological manifestations in particular [16]. The systemic inflammatory response seen in adults is much rarer in the pediatric population [17]. However, long-COVID may well be an under-recognized phenomenon [18]. The challenge of recognizing the clinical characteristics of COVID-19 among other neonatal diseases is apparent [19]. Some recognized manifestations of SARS-CoV-2 infection in children include stroke [20], seizures or seizure-like activities [21-23], headaches [21], dizziness [21], meningoencephalitis [21,24-26], and opsoclonus [27]. Rare neurological complications include intracranial hemorrhage, cranial neuropathies, Guillain-Barré syndrome, and visual impairments [28]. Many of the neurologic manifestations are observed in association with multisystem inflammatory syndrome in children [29]. The underlying pathophysiology driving these clinical manifestations of SARS-CoV-2 infection remain to be fully elucidated, although immune dysregulation and autoreactivity have been shown to correlate with disease severity in children [30], and the use of blood biomarkers for the diagnosis of brain injury in patients with COVID-19 have been proposed [31]. Neuroimaging performed in children with SARS-CoV-2 infection most commonly shows postinfectious immune-mediated acute disseminated encephalomyelitis-like changes in the brain, myelitis, and neural enhancement [32]. Focal cerebral arteriopathy [33] has also been identified. In the neonatal population, diffusion restriction [34] and cerebral white matter injury [35] have been described.

The effect of SARS-CoV-2 infection specifically in neonates has been relatively understudied to date [36,37]. There is evidence to indicate that most infected neonates are asymptomatic or experience only mild disease [38], although risks of direct and indirect adverse health outcomes have been identified [39] and the risk of neonatal brain injury from SARS-CoV-2 has been highlighted in the literature [40]. There is also evidence that SARS-CoV-2 infection is higher in premature neonates, and this appears to be due to premature delivery secondary to maternal reasons as opposed to an increase in spontaneous preterm labour [41].

Neurodevelopmental outcomes of neonates infected with SARS-CoV-2 are of particular interest given the potential implications for the child’s long-term prognosis and functional outcomes. There has been little previous work in this area to date [42]. A study has shown that in utero exposure to maternal SARS-CoV-2 infection is not associated with neurodevelopmental outcome at age 6 months [11], and another study has shown that newborns exposed in utero to SARS-CoV-2 have largely normal neurological development in the first few months of life. There is an ongoing multicenter observational study currently recruiting that also aims to review neurodevelopmental outcomes at six months [43], although longer-term follow-up is required to provide a more accurate picture. Telehealth may provide a useful way to monitor the long-term neurodevelopmental outcomes of neonates who develop SARS-CoV-2 infection [44].

There are several limitations to the current study. First, the control population were a historical cohort, and thus their data may be incomplete. Second, the sample size was relatively small. Third, there were some confounders that we were unable to control for.

In conclusion, our study demonstrates no difference in neurodevelopmental outcomes in children infected with SARS-CoV-2 infection compared to a historical control group. However, additional long-term follow-up is required to ensure this remains the case after 18 months. Furthermore, the neurodevelopmental outcomes are likely to be largely normal because most neonates are asymptomatic or have mild symptoms; neonates with severe COVID-19 or neurological involvement are likely to be more at risk of neurodevelopmental impairment.

## Data Availability

All data produced in the present study are available upon reasonable request to the authors

## Abbreviations

BSID-III: Bayley Scales of Infant and Toddler Development-3rd Edition
COVID-19: coronavirus disease 2019
GMFCS: Gross Motor Function Classification System
IQR: interquartile range
MIS-C: multisystem inflammatory syndrome in children
SARS-CoV-2: severe acute respiratory syndrome coronavirus-2
SD: standard deviation

## Acknowledgements

We would like to thank parents and children participating in the study and the respective register holders; Pediatric COVID-19 Kuwait National Registry.

